# Aneurysm Analysis Using Deep Learning

**DOI:** 10.1101/2025.06.19.25328680

**Authors:** Alireza Bagheri Rajeoni, Breanna Pederson, Susan M. Lessner, Homayoun Valafar

## Abstract

Precise aneurysm volume measurement offers a transformative edge for risk assessment and treatment planning in clinical settings. Currently, clinical assessments rely heavily on manual review of medical imaging, a process that is time-consuming and prone to inter-observer variability. The widely accepted standard-of-care primarily focuses on measuring aneurysm diameter at its widest point, providing a limited perspective on aneurysm morphology and lacking efficient methods to measure aneurysm volumes. Yet, volume measurement can offer deeper insight into aneurysm progression and severity. In this study, we propose an automated approach that leverages the strengths of pre-trained neural networks and expert systems to delineate aneurysm boundaries and compute volumes on an unannotated dataset from 60 patients. The dataset includes slice-level start/end annotations for aneurysm but no pixel-wise aorta segmentations. Our method utilizes a pre-trained UNet to automatically locate the aorta, employs SAM2 to track the aorta through vascular irregularities such as aneurysms down to the iliac bifurcation, and finally uses a Long Short-Term Memory (LSTM) network or expert system to identify the beginning and end points of the aneurysm within the aorta. Despite no manual aorta segmentation, our approach achieves promising accuracy, predicting the aneurysm start point with an R^2^ score of 71%, the end point with an R^2^ score of 76%, and the volume with an R^2^ score of 92%. This technique has the potential to facilitate large-scale aneurysm analysis and improve clinical decision-making by reducing dependence on annotated datasets.

## 1. Introduction

Abdominal aortic aneurysms (AAAs) are defined as abnormal dilations of the abdominal aorta measuring 3.0 cm or more in diameter. This condition arises when the aortic wall weakens, leading to a localized bulging that can progressively enlarge over time. The prevalence of AAAs increases with age, particularly affecting individuals over 60 years old, and is more common in men and those with a history of smoking [1].

The risk of aneurysm rupture increases with size, but even aneurysms below the accepted clinical intervention cut-off of 5.5 cm can pose a significant danger. Aneurysms between 4.5 and 4.9 cm have shown an average expansion rate of 0.7 cm per year, and ruptures have been reported in patients with aneurysms as small as 5.0 to 5.6 cm [2]. These findings highlight the importance of close monitoring and early intervention, even for aneurysms below the traditional surgical threshold. Traditionally, the maximum transverse diameter (MTD) has been the standard measurement for evaluating AAAs [3] [4]. However, this linear measurement may not fully capture the complex three-dimensional nature of aneurysms. Relying on a single diameter measurement can overlook shape irregularities, aneurysm length/tortuosity, and other geometric factors. In fact, diameter is considered a “rough and ready” measure and shows poor ability to detect certain shape changes or small growth increments, underscoring its limitations in reflecting the aneurysm’s true three-dimensional expansion [4], [5]. In addition to diameter, advanced biomechanical criteria, such as stress and strain analysis, have been proposed to enhance AAA evaluation and rupture risk assessment. Peak wall stress (PWS), a biomechanical parameter, quantifies the maximum stress on the aneurysm wall under blood pressure, and studies show that it is a more reliable rupture risk indicator than diameter, with higher PWS observed in ruptured or symptomatic AAAs compared to electively repaired ones [6], [7]. Finite element analysis (FEA) uses patient-specific 3D geometry from CT scans, incorporating wall thickness, material properties, and pressure to compute stress distributions, revealing high-stress regions prone to rupture [6], [8]. Strain analysis, measuring deformation, complements this by indicating how the wall stretches under load, with 3D ultrasound and CT-based methods tracking wall motion across the cardiac cycle to produce strain maps that highlight areas of excessive deformation, signaling rupture risk [9]. The rupture potential index (RPI), a ratio of wall stress to wall strength, integrates 3D geometry from CT scans, FEA-derived stress, and estimated wall strength, showing elevated values in symptomatic and ruptured AAAs, offering a nuanced prediction of rupture risk [6], [10]. However, these methods face challenges in clinical adoption, requiring accurate 3D reconstructions, precise wall thickness measurements (limited by CT resolution), and patient-specific material properties, which are hard to obtain non-invasively [8].

Recent studies suggest that volumetric assessments provide a more comprehensive evaluation of aneurysm morphology and may better predict clinical outcomes. For example, a systematic review indicated that volume measurements could offer a more accurate assessment of aneurysm growth compared to diameter measurements [4]. Large-scale analyses from the M2S database reveal that post-repair sac enlargement occurs in up to 40% of thoracic and abdominal aneurysms, a risk strongly tied to preoperative anatomy that diameter measurements often underestimate [11]. By accounting for the entire aneurysm sac, volume captures changes along the length and width of the aneurysm that a single diameter cannot. Accordingly, adding volume measurements to diameter has been shown to yield additional information on AAA growth, improving the characterization of expansion over time [3], [4]. Despite these findings, volumetric analysis has not been widely adopted in clinical practice, partly due to the lack of standardized measurement protocols and the labor-intensive nature of manual volume calculations. Measuring volume often requires manual or semi-automated image segmentation, which demands specialized software and expertise and can take significant time, making it impractical for busy clinics [4] [12].

Advancements in CT technology (e.g. helical CT and multi-slice scanners) have made imaging both fast and high-resolution. Helical CT permits much quicker scan times (on the order of seconds, often in a single breath-hold) than older CT modalities, virtually eliminating motion artifacts. These CTA techniques provide a combination of speed and detail, producing rapid, high-definition images of the AAA and its surrounding structures. Such 3D CTA images clearly show aneurysm size, shape, and involvement of branch vessels, offering an unprecedented anatomical overview compared to traditional 2D angiography. Notably, CTA is also less invasive than catheter angiography and simultaneously allows for evaluation of other abdominal pathology during the same scan [13]. However, the interpretation of these images often relies on manual analysis by radiologists, which is time-consuming and subject to inter-observer variability.

To address these challenges, there is a growing interest in developing automated methods for aneurysm detection and measurement. Artificial Intelligence (AI) and deep learning algorithms have shown promise in healthcare, specifically accurately analyzing vascular structures [14], [15], [16] [17], [18], [19], [20], [21]. For instance, recent research demonstrated the use of deep learning techniques to automate the measurement of vascular calcifications, highlighting the potential applicability of such methods to AAA assessment [22], [23], [24]. Automated approaches can potentially reduce the workload of clinicians, minimize human error, and provide consistent measurements, thereby enhancing the reliability of aneurysm evaluation.

Despite these technological advancements, the implementation of automated systems in clinical practice faces several obstacles. A significant limitation is the scarcity of large, annotated datasets required to train robust AI models. Thousands of labeled medical images, along with corresponding diagnostic and prognostic information, are essential to develop models that generalize well across diverse patient populations. Also, the collection and management of such extensive datasets are hindered by privacy concerns, high annotation costs, and the need for scalable data processing infrastructures [25], [26]. The current lack of such datasets hampers the progress of AI-driven diagnostic solutions and underscores the need for collaborative efforts to create comprehensive medical imaging repositories.

This study aims to detect and quantify aneurysms by developing an automated method capable of accurately identifying aneurysms where there is no segmentation label data available. By enabling comprehensive and reliable evaluation and analyzing the progression of the AAAs, this approach has the potential to enhance clinical decision-making, support timely interventions, and reduce the reliance on time-consuming and variable manual assessments.

In summary, while traditional methods of aneurysm assessment have relied heavily on manual measurements of maximum diameter, emerging evidence supports volumetric analysis as having the potential to further improve accurate evaluation of aneurysm size and growth, complementing traditional methods. The integration of advanced imaging techniques and automated measurement tools holds promise for improving the detection, monitoring, and treatment planning of AAAs. However, the successful implementation of these technologies in clinical settings will require overcoming challenges related to data availability, standardization of measurement protocols, and validation of AI algorithms. https://github.com/pip-alireza/automated_aneurysm_analysis/ hosts the resources for this paper.

## 2. Materials and Methods

### 2.1. Data Description and Annotation

The dataset used in this study consists of de-identified computed tomographic angiography (CTA) scans from 60 patients with AAAs, obtained from the M2S Vascular Imaging Database (West Lebanon, NH, USA). M2S provides imaging analysis services and serves as the core lab for multiple aneurysm research studies [11], [27]. For this work, they supplied fully de-identified DICOM series, including both preoperative and postoperative scans. All study design, analysis, and interpretation were conducted independently of M2S.

To generate ground truth for aneurysm boundaries, patient scans were manually annotated by an expert in arterial mechanics. The manual annotation process involved identifying the specific slice indices corresponding to the onset and conclusion of the aneurysm, based on morphological changes, primarily variations in cross-sectional diameter, as shown with number 1 and 2 in Figure 1.

**Figure 1.**
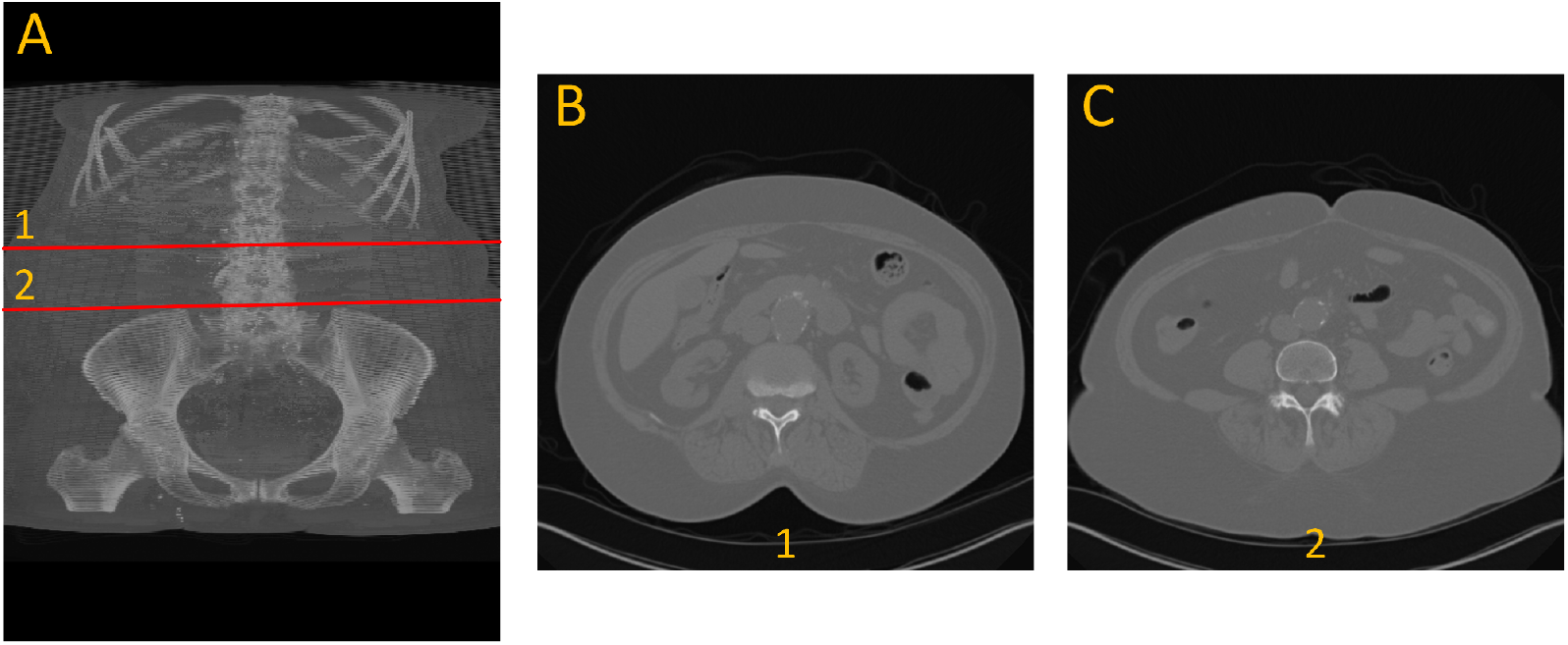
Example of the dataset. (A) 3D view front with annotation of the beginning (1) and end (2) of aneurysm. (B) cross section view of the beginning of the aneurysm. (C) cross section view of the end of aneurysm.

### 2.2. Data Preprocessing

CTA images were first converted from DICOM to TIFF files and normalized to an 8-bit grayscale format using linear contrast stretching (range: 10–245) and numpy library. To be compatible with segmentation models, grayscale TIFF images were duplicated across three channels. For the LSTM model, the segmented pixel counts per slice (output from SAM2) were extracted and formatted into 1D sequences per patient scan. This sequence of information can be used for visual inspection of the aneurysm and as the primary information to be used for the development of aneurysm detection method.

### 2.3. Deep Learning Approach for Automated Aneurysm Analysis

We pursued two complementary strategies for aneurysm detection. The first relied on transfer learning from our earlier arterial-system segmentation studies [18], [22], [19]. By re-deploying a UNet trained solely on normal anatomy, we could screen new scans that lack aneurysm annotations: In normal slices the model segments the vessel as expected, but it stops segmenting whenever the aortic wall becomes irregular, effectively flagging those gaps as potential disease. Interpreting this absence of segmentation as an aneurysm cue, and by applying simple rule-based logic, we could predict the presence of the aneurysm, although this transfer-learning approach does not provide volume measurements.

The second and main strategy consisted of utilizing an ensemble of tools or multi-system approach. This approach utilized the Segment Anything Model 2 (SAM2) developed by Meta [28] in conjunction with our previous UNet model [19], [22]. This approach starts with UNet providing the initial segmentation of the aorta, then SAM2 tracks the vessel through every slice, even across irregular regions. Boundaries are identified either by a rule-based expert system or by a bidirectional LSTM that learns temporal patterns in the pixel-count signal. Once boundaries are set, slice integration provides aneurysm volume, and linear interpolation between boundary slices estimates the normal baseline for enlargement quantification. In testing, the expert-system variant delivered the most accurate volumes and a strong Dice overlap, while the LSTM version achieved slightly lower but still solid agreement. Compared with the transfer-learning screen, this pipeline adds boundary placement and volumetric measurements. The architecture comprises the following components:

#### Aorta Localization (UNet)

A pre-trained UNet model with an encoder-decoder structure and a ResNet-34 backbone, segments and localizes the aorta from CTA images [22], [29]. Despite its limitations when encountering pathological variations like AAA, the UNet reliably identifies normal regions of the thoracic aorta, providing initial slice numbers and center coordinates that serve as input prompts for subsequent models.

#### Aorta Tracking (SAM2)

The Segment Anything Model 2 (SAM2) employs a transformer-based streaming memory mechanism to track the aorta continuously from the prompted region down to the iliac bifurcation. It begins with a prompt provided by the UNet model, which identifies a region of interest within the aorta. SAM2 then refines this segmentation by searching for regions exhibiting similar characteristics to the prompted area, effectively accounting for morphological changes along the vessel. The model outputs both segmentation masks and the count of segmented pixels per slice, facilitating detailed analysis of the aorta’s structure.

#### Aneurysm Boundary Identification

Two complementary approaches are used:

##### 1. LSTM-Based Aneurysm Detection

To identify the beginning and end points of the aneurysm within the aorta, we developed a Long Short-Term Memory (LSTM) model that analyzes sequential changes in aortic cross-sectional area. From the SAM2 outputs, we collected the number of segmented pixels per slice, effectively forming a 1D sequence representing area changes across axial slices. The model architecture comprises:

a. Two **stacked bidirectional LSTM layers**, each with 600 hidden units, designed to capture forward and backward temporal dependencies in the sequence.
b. A series of **fully connected layers** with ReLU activation, mapping LSTM outputs to slice-wise classification scores across 200 output neurons, covering the region from the thoracic aorta to the iliac bifurcation.
c. A **sigmoid-activated output layer**, producing per-slice probabilities between 0 and 1.
d. A fixed **threshold** to classify each slice as aneurysmal or normal, enabling start and end slice identification.

##### 2. Expert Rule-Based System

This component implements a sliding-window analysis that computes the average segmented pixel count over a fixed number of preceding slices to the output of SAM2. If a slice’s value exceeds 120% (for the start) or drops below 80% (for the end) of the running average consistently across the window, it is flagged as an anomaly corresponding to aneurysm boundaries. We performed a grid search to optimize the window size and the upper and lower threshold parameters for best performance.

#### 2.4. Evaluation Metrics

Because no single statistic captures every aspect of model performance, we evaluated our pipelines with a suite of complementary measures. The UNet and SAM2 components were adopted exactly as trained in prior work, UNet on non-aneurysmal aorta [19], [22] and SAM2 as distributed by Meta [28], whereas the LSTM that refines aneurysm boundaries was trained from scratch on our dataset. Five-fold cross-validation ensured generalisability: in each fold, 80% of the patients were used to train the LSTM and 20% to test it, with strict patient-level separation to avoid data leakage. Training relied on the Adam optimiser, learning-rate 0.0003, batch-size 10, and binary-cross-entropy loss combined with a Jaccard term; early stopping on validation loss curtailed over-fitting.

##### Overlap quality

The spatial accuracy of each predicted boundary was assessed using the Dice coefficient, as defined in Equation (1). The Dice score, also known as the F1 score, is computed by doubling the intersection of the predicted and ground truth masks and dividing by the sum of their areas. In this context, it evaluates how well the predicted boundary aligns with the annotated boundary. Specifically, the Dice score uses true positives (TP), representing the correctly predicted overlapping regions between the predicted and ground truth masks, false positives (FP), indicating regions incorrectly predicted as part of the boundary, and false negatives (FN), denoting regions missed by the prediction but present in the ground truth. The Dice score ranges from 0 (no overlap) to 1 (perfect match), providing a clear measure of how well the predicted mask aligns with the ground truth.

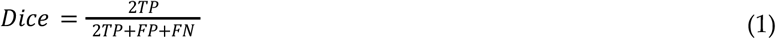

##### Boundary accuracy

Start and end slice predictions were compared with ground truth using the coefficient of determination (*R*^2^), mean absolute error (MAE), and mean squared error (MSE) to evaluate per-case deviations. The *R*^2^ metric, shown in Equation (2), represents the proportion of variance in the ground-truth values that is explained by the predicted values. It is calculated using the Residual Sum of Squares (RSS), which measures the sum of squared differences between the predicted and actual values, and the Total Sum of Squares (TSS), which quantifies the total variance in the ground truth data. A higher R^2^ value indicates better predictive accuracy, with 1 representing a perfect fit and 0 indicating no explanatory power.

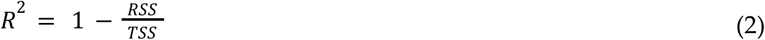

In addition, Equation (3) defines MAE, which quantifies the average absolute difference between predicted and actual slice indices. And Equation (4) defines MSE, which captures the average squared difference between predicted and true values, penalizing larger errors more strongly:

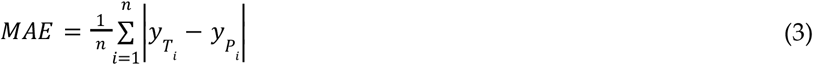

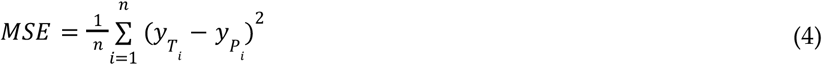

Lower values imply higher quantitative fidelity. In these equations, 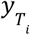 represents the true value for slice *i*, 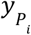 represents the predicted value for slice *i*, and *n* is the number of samples.

##### Volumetric agreement

For cases where SAM2 produced a complete aneurysm mask, the aneurysm volume was computed by integrating the segmented pixel values across slices. Agreement with manual volume was assessed using the R^2^ metric.

##### Training diagnostics

During optimization of the LSTM model, we monitored binary cross entropy (BCE), and tracked the score on a held-out validation subset after each epoch. Once training and validation losses converged and further epochs failed to improve score, the model with the best score was frozen and evaluated on the untouched test folds. Binary cross entropy is expressed by Equation (5).

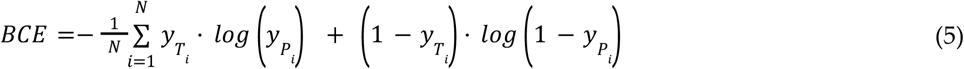

##### Volumetric agreement

We treat the sum of segmented pixels within the aneurysm boundaries as a proportional volume surrogate. If the predicted start and end slices are *s* and *e, P*_*i*_ denotes the number of segmented pixels in slice *i*. The surrogate volume is computed by Equation (6).

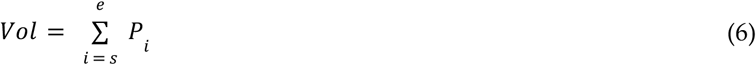

This unit-less figure scales linearly with true physical volume. Although it does not yield an absolute volume in cubic millimetres and ignores wall thickness, it is sufficient for primary algorithm validation, allowing consistent comparison with pixel totals when aneurysm boundaries are identified manually. Agreement between automated and manual surrogate volumes is reported with *R*^2^; a value close to 1 indicates that the model captures nearly all variability in the reference measurements despite the absence of geometric calibration. By combining Dice, MAE, MSE and *R*^2^ for both boundary indices and volumes, we obtained a comprehensive view of each pipeline’s ability to locate, delineate and quantify abdominal aortic aneurysms.

#### 2.5. Prior Work and Motivation

Our earlier work established the utility of deep learning for vascular analysis: a UNet-based system achieved an 83.4 % Dice score for segmenting the arterial tree from the descending thoracic aorta to the knees, produced automated calcification scores that correlated highly with manual assessments, *R*^2^ = 0.978, and yielded a MAPE of just 9.5 % [22]; a subsequent transformer-based model, TransONet, raised segmentation performance to 93.5 % Dice from the thoracic aorta to the iliac bifurcation and maintained 80.64 % Dice down to the knees [18]. These successes motivated us to pursue aneurysm detection and volume quantification using similar approaches.

## 3. Results and Discussion

### 3.1. Using UNet Failure for Aneurysm Boundary Identification

We began by applying our previously trained UNet model [22], trained on normal vascular system data, to identify aneurysms indirectly via segmentation failure. The model failed to segment the aorta in regions with anomalies. In essence, encountering an “unseen” pathological structure led the model to produce no segmentations. However, we observed that the absence of segmentation in anomalous slices effectively acted as a binary marker distinguishing “normal” from “abnormal” vascular systems. We developed an expert rule that interprets segmentation failure as an indicator of pathology. By maintaining a rolling average of segmented pixel counts across four consecutive slices, we established a baseline for normal aortic segmentation. Any slice where the segmentation dropped below 50% or exceeded 140% of this baseline was flagged as anomalous. If these deviations persisted across four consecutive slices, the first flagged slice was marked as the aneurysm start, while the last was designated as the end of the aneurysm. These threshold parameters and window size were optimized through a grid search to achieve the best performance. We tested this rule-based approach on 33 patient scans: 16 non-aneurysm cases sourced from our earlier study [22] and 17 cases with AAAs. The resulting confusion matrix is shown in Table 1. The expert system correctly identified 14 out of 16 non-aneurysm cases and 15 out of 17 aneurysm cases, yielding an overall accuracy of 87.9%. Precision, recall, and F1-score were all 88.2%, while specificity was 87.5%. These results demonstrate that segmentation failure in a UNet trained only on normal anatomy can serve as a reliable binary classifier for vascular abnormalities. Figure 2 (A) shows how the UNet fails to segment the aneurysmal region, while (B) illustrates successful segmentation when SAM2 is used in conjunction with UNet.

**Table 1.** Confusion matrix for the UNet + expert-rule classifier.

|  | Predicted Normal | Predicted Aneurysm |
| --- | --- | --- |
| Actual Normal | 14 | 2 |
| Actual Aneurysm | 2 | 15 |

**Figure 2.**
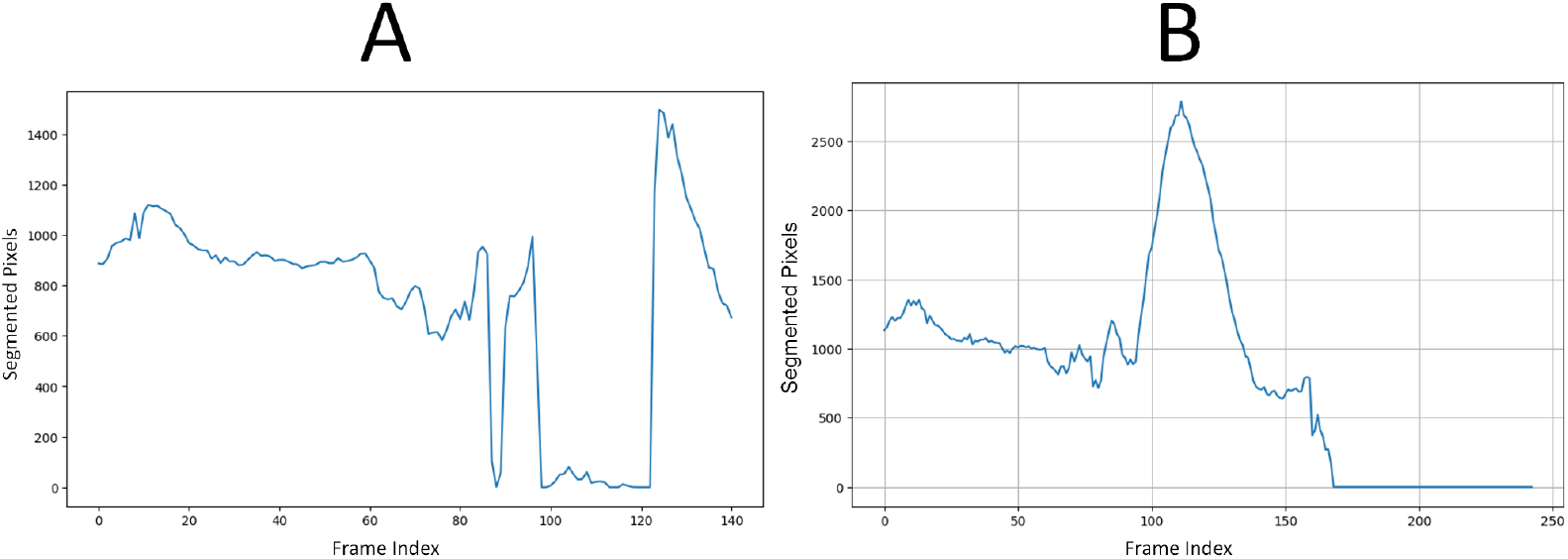
Vascular system segmentation with (A) UNet alone, versus (B) UNet + SAM2 on the same patient scan. The standalone UNet ceases to generate a mask in the aneurysmal region, visible between slices ~95 and ~125, whereas the UNet + SAM2 pipeline preserves a continuous aortic segmentation through the aneurysmal region.

### 3.2. Aneurysm Localization and Segmentation

Despite its limitations, the UNet accurately localized normal aortic slices. We extracted the first slice with successful segmentation and established the center coordinates (x, y) of the aorta. These served as point prompts for the SAM2 model. Using point prompts from UNet, the SAM2 model tracked the aorta throughout the entire scan. It adapted its segmentation to changes in aortic shape, including aneurysmal regions. The result is a per-slice segmentation mask and a count of segmented pixels representing cross-sectional area.

Figure 3 illustrates this full pipeline. Panel A shows the original CTA scan, while B displays the complete aorta mask generated by SAM2 after propagation over the input image. C shows the segmented mask from SAM2 output and finally, panel D highlights the aneurysm region using the manual annotation, with red indicating normal aorta segment and blue denoting the aneurysm segment. This visualization demonstrates how UNet and SAM2 operate in tandem: UNet anchors the aorta in normal regions, and SAM2 tracks it seamlessly across the aneurysm segments for full-volume segmentation. Figure 4 provides a cross-sectional view at the aneurysm’s start and end boundary, further illustrating the model’s performance.

**Figure 3.**
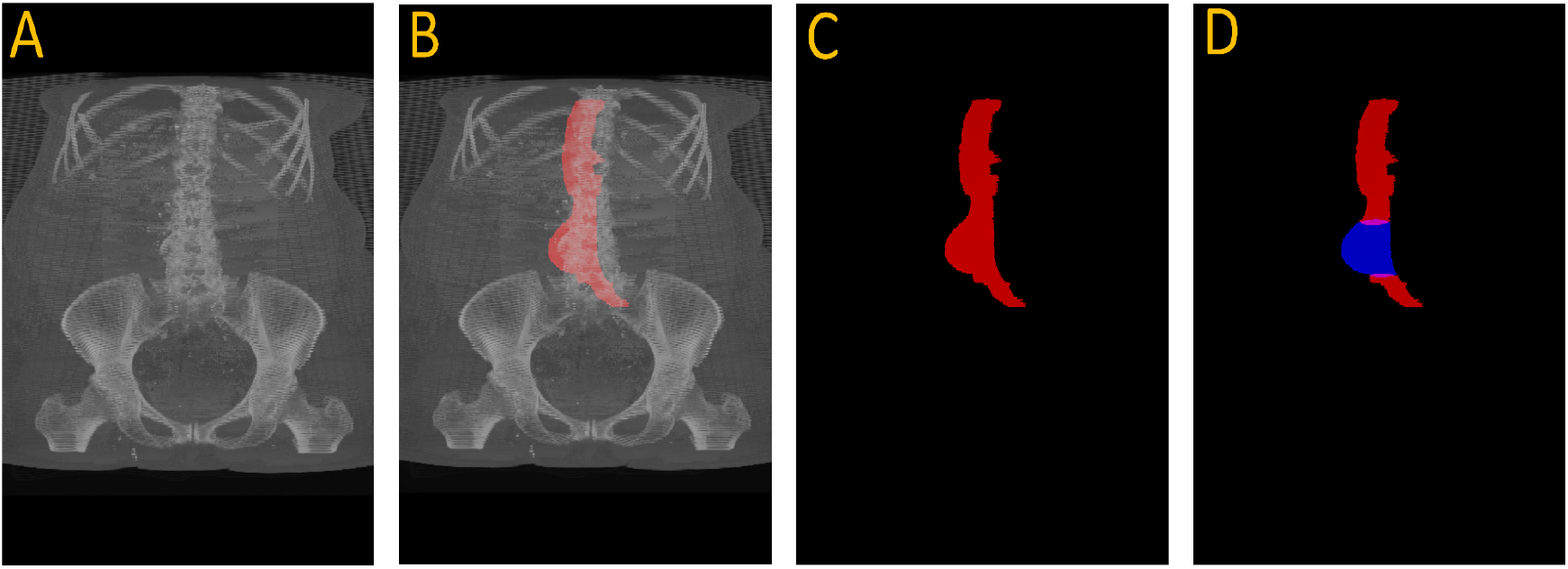
Full aneurysm segmentation pipeline. (A) Original CTA scan; (B) SAM2-generated aorta mask overlaid on the scan after propagation; (C) Binary segmentation mask produced by SAM2; (D) Manual annotation showing normal aorta (red) and AAA region (blue). The sequence shows how the proposed approach extracts an aneurysm despite having no aneurysm labels during training.

**Figure 4.**
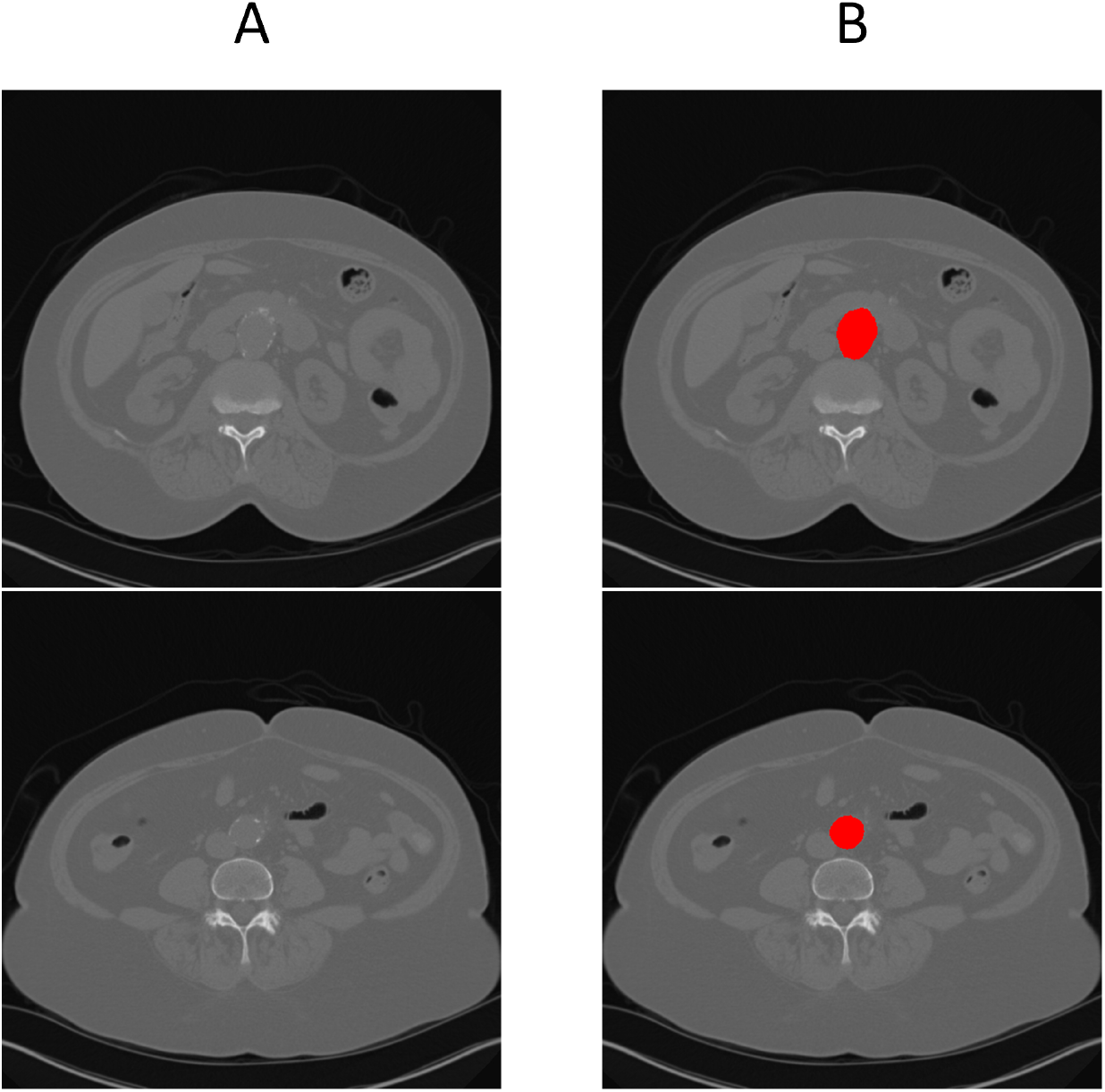
Cross-sectional view of SAM2’s segmentation performance at the start and end boundaries of an aneurysm (refer to Figure 1 for anatomical reference).

To enable full automation of aneurysm analysis, including detection, boundary localisation, and volume estimation, we developed two downstream boundary detection strategies: a rule-based expert system and a learnable LSTM model. Both operate directly on the pixel-count sequences output by SAM2, allowing automated identification of aneurysm start and end boundaries without the need for manual annotation.

#### 3.2.1. Results of UNet + SAM2 + LSTM (USL) Approach

##### 3.2.1.1. Training and Validation Loss

The bidirectional LSTM was trained for 1000 epochs on slice-wise pixel-count sequences, using five-fold cross-validation with an 80 / 20 split between training and validation patients. Both loss curves dropped sharply in the early epochs and then flattened at almost identical values; the validation trajectory never diverged from the training trajectory, indicating minimal over-fitting and good generalisability across folds. Figure 5 illustrates the training and validation loss for one of the folds. As we can see from the figure, the model starts to over fit on the training around 500 epochs and validation loss did not improve after that. Figure 5 shows LSTM training versus validation loss over 1,000 epochs. Both curves drop sharply during the first 50–100 epochs, indicating rapid learning, and then flatten to a stable plateau. Training loss settles near 0.15 and validation loss near 0.22.

**Figure 5.**
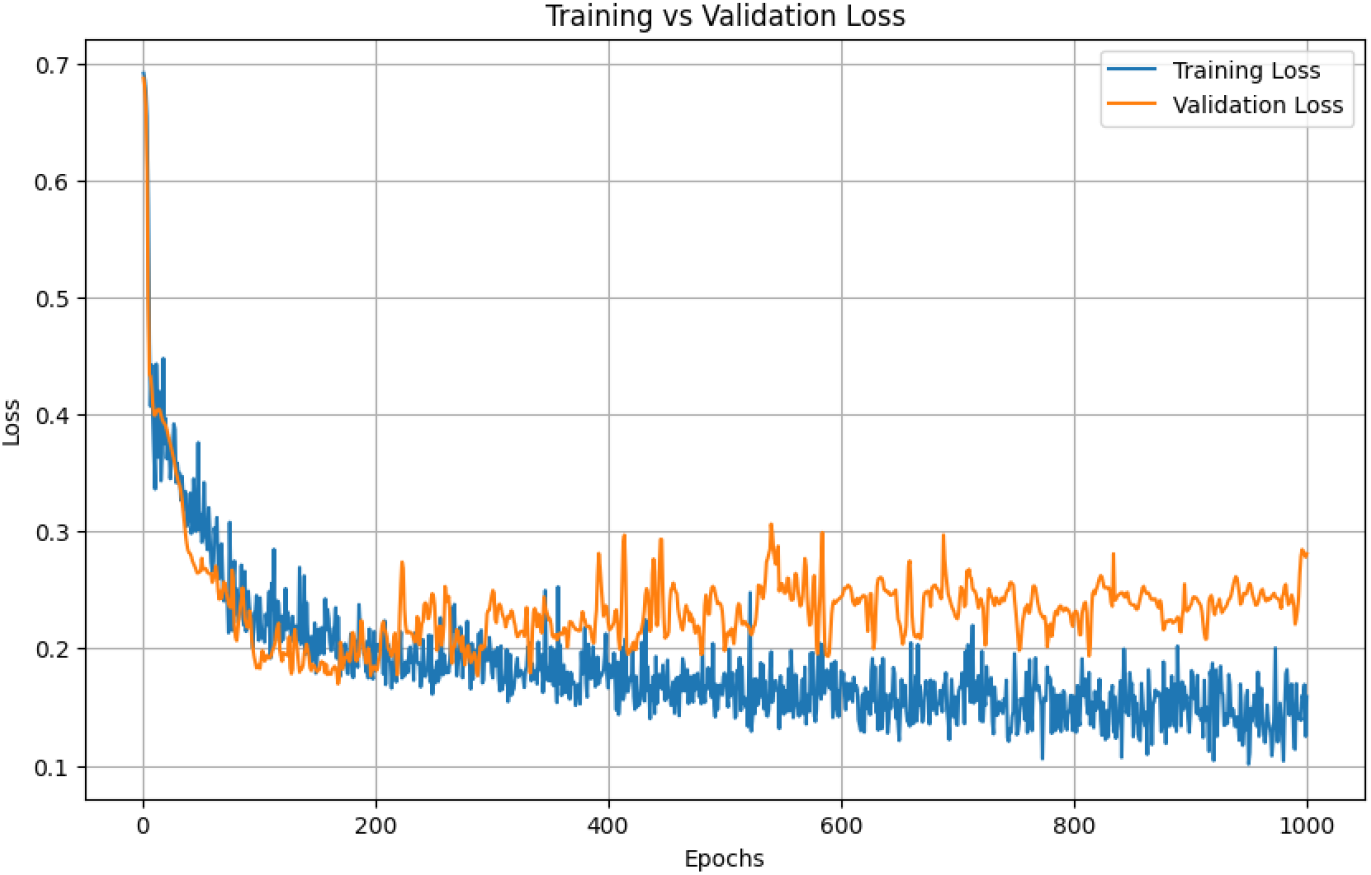
Training and validation loss for LSTM.

##### 3.2.1.2. Boundary-Prediction Accuracy

Predicted start and end slices were compared with ground truth. Across the full cross-validation the aneurysm segment achieved an average Dice overlap of 65 %, confirming that the pipeline provides clinically acceptable boundary localisation. Figure 6 illustrates the performance of USL on predicting the boundary in a subset of testing data. The Dice score reflects the degree of overlap between the red and green regions in Figure 6, with higher overlap indicating better segmentation accuracy.

**Figure 6.**
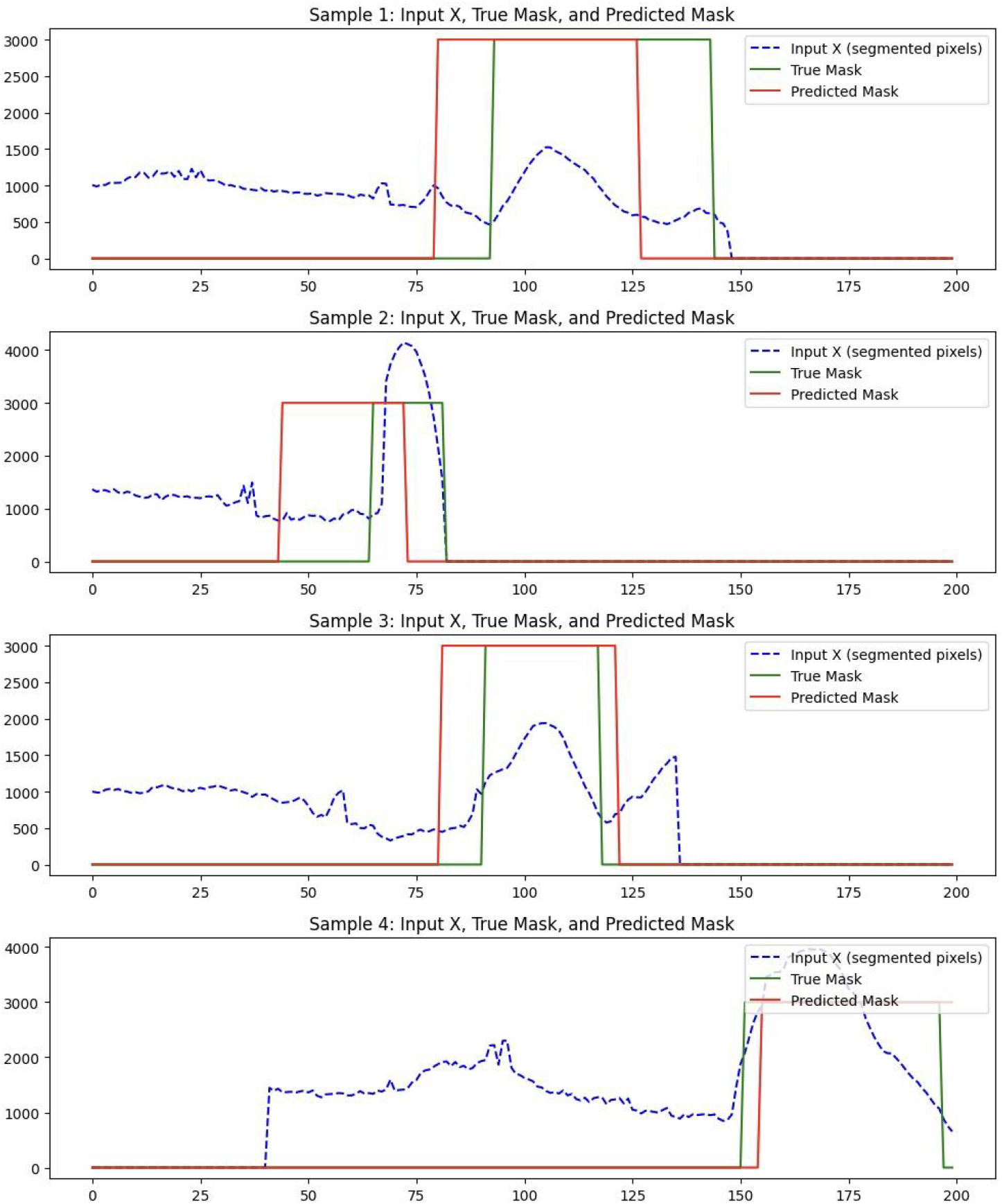
USL performance on predicting the boundary of aneurysm on 4 samples.

##### 3.2.1.3. Volume-Estimation Accuracy

Integrating the SAM2 mask pixel counts between the LSTM-predicted boundaries produced a surrogate lumen volume that showed moderate agreement with manual measurements, achieving an *R*^2^ of 0.57 as shown in Figure 7. This indicates the automated pipeline captures just over half of the variance observed in expert-derived volumes, reflecting useful, but not yet optimal quantitative accuracy.

**Figure 7.**
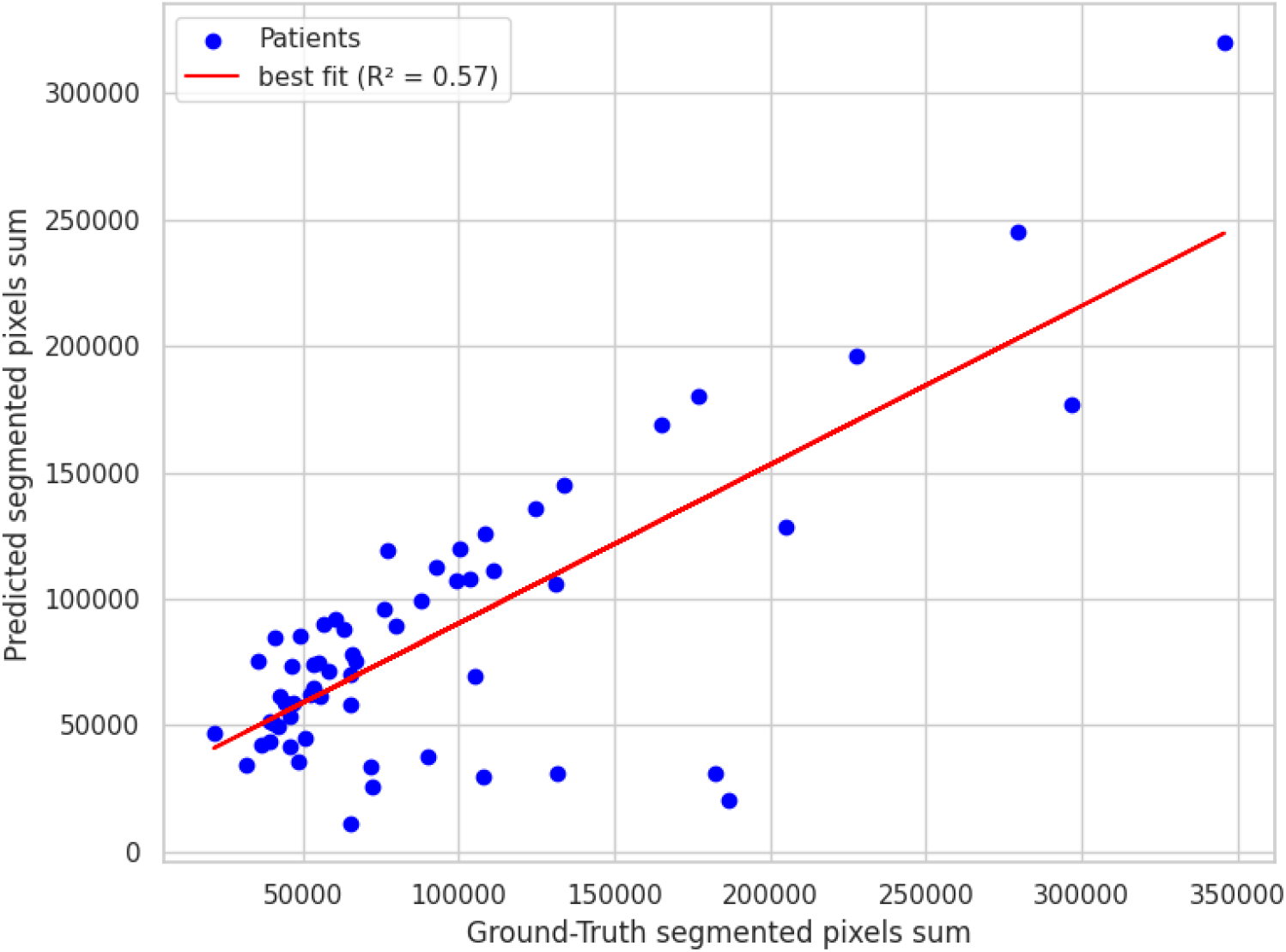
USL performance on predicting the aneurysm volume compared to ground truth.

#### 3.2.2 Results of UNet + SAM2 + Expert (USE) Approach

In an alternative pipeline, we employed an expert system in place of the LSTM for aneurysm boundary detection. The rule consists of scanning through the number of segmented pixels for each cross-sectional slice and detecting significant rises and drops over a sliding window of four slices. Once the largest “abnormality” window is identified, the start and end of that region are deemed to be the aneurysm boundaries. The results are summarized below. We used grid search to find the optimum values for the window size, upper and lower threshold.

##### 3.2.2.1. Boundary prediction accuracy of expert system

We measured the boundary prediction performance of the expert system against the ground truth for the beginning and end of the aneurysm. While minor discrepancies were observed, the expert system outperformed the USL approach. It achieved an average Dice score of 78%, indicating strong spatial overlap with the region annotated manually. Figure 8 illustrates the performance of USE on predicting the boundary in a subset of testing data.

**Figure 8.**
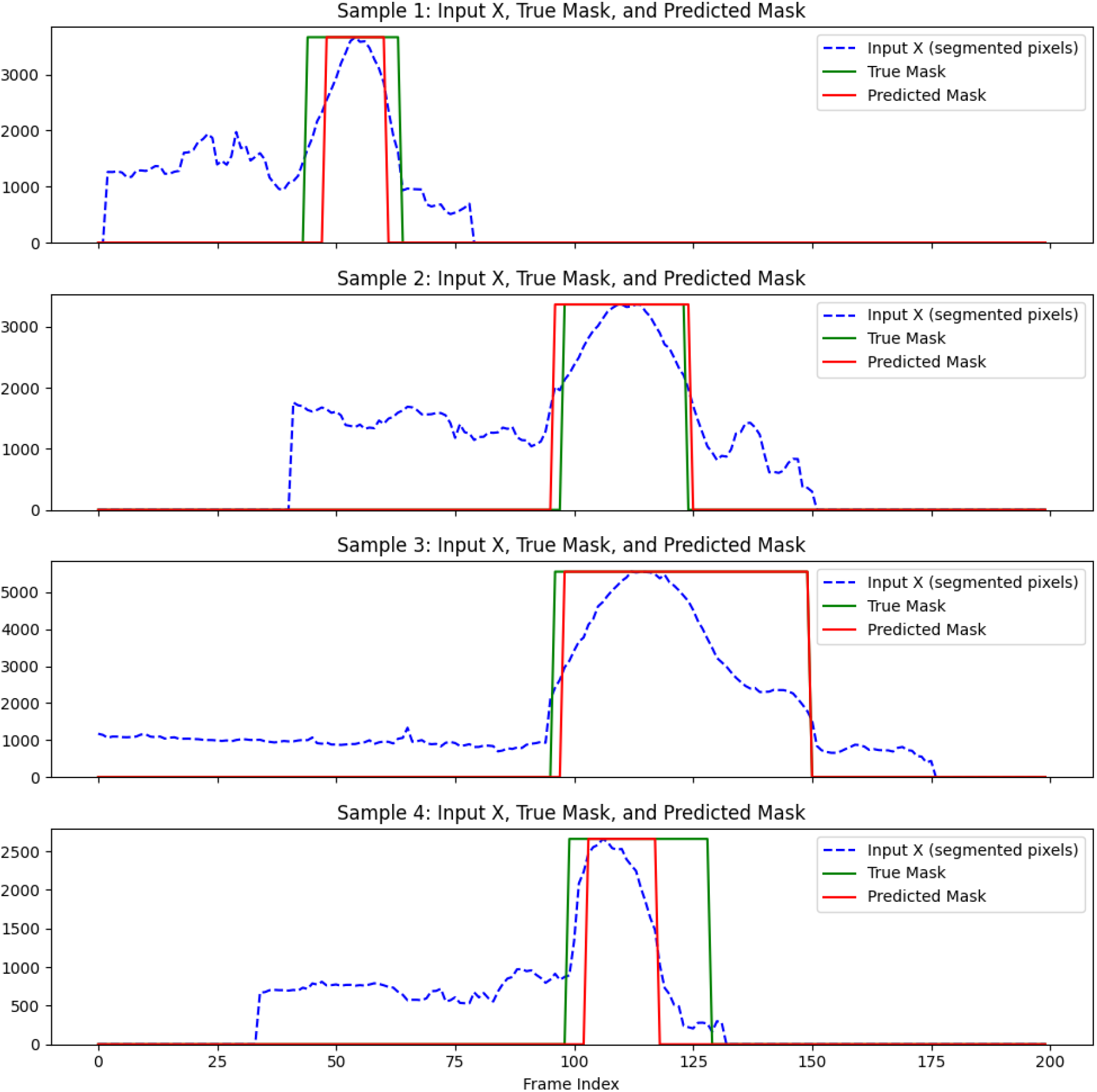
USE performance on predicting the boundary of aneurysm compared to ground truth.

##### 3.2.2.2 Volume Estimation Accuracy of expert system

The scatter plot shown in Figure 9 shows a strong correlation in volume estimation, with an R-squared value of 0.92 in segmented pixel counts between the expert system and manual boundary annotations. This indicates that the expert system aligns well with manual annotations, demonstrating a robust capacity for accurate volume measurement.

**Figure 9.**
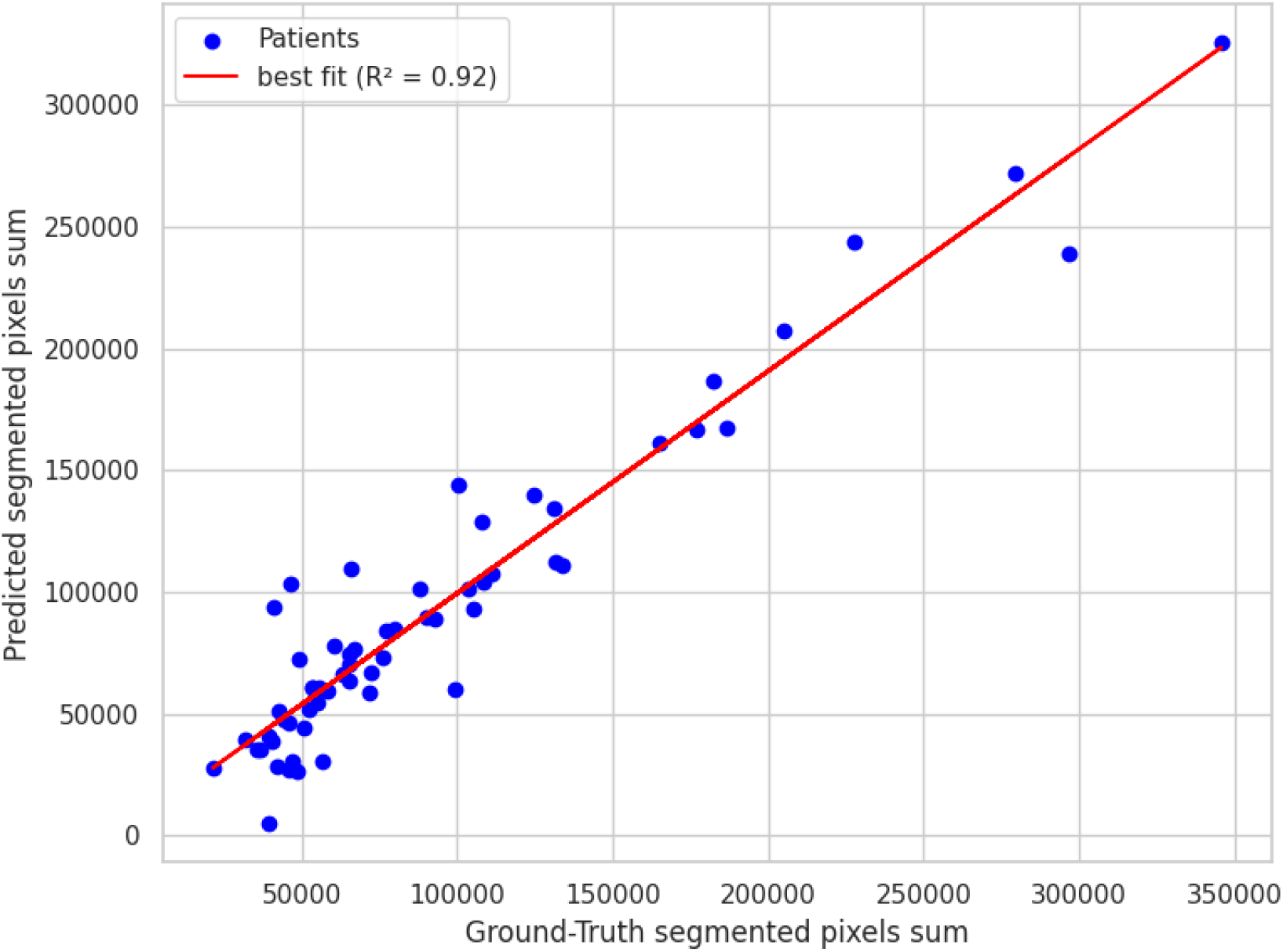
USE performance on predicting the aneurysm volume compared to ground truth..

### 3.3. Comparison of Methods

Table 2 illustrates a side-by-side look at the start and end boundaries detection performance, using R^2^, MAE, and MSE, and the Average Dice Score across two different pipelines:

**Table 2.** Performance comparison between USL and USE across multiple evaluation metrics.

| Model | $R^2$ Start | $R^2$ End | MAE Start | MAE End | MSE Start | MSE End | Dice | $R^2$ Volume |
| --- | --- | --- | --- | --- | --- | --- | --- | --- |
| USL | 0.183 | 0.340 | 17.750 | 13.017 | 449.31 | 410.18 | 0.65 | 0.572 |
| USE | 0.714 | 0.759 | 7.667 | 8.26 | 157.36 | 148.96 | 0.78 | 0.916 |

Figures 10 and 11 summarize the comparative performance of the two pipelines USL and USE across six sub-panels. Figure 10 (A) displays the R^2^ values for the start boundary, end boundary, and volume, immediately highlighting USE’s superior explanatory power at all three targets. Figure 10 (B) and (C) contrasts absolute error for the start and end indices, where the noticeably shorter USE bars confirm its finer boundary localisation. Figure 10 (D) displays histogram plots of mean absolute errors, while Figure 10 (E) shows the mean-squared errors; in both, USE has better performance delineating the boundary than USL. Figure 11 blends a violin plot with an embedded box plot to reveal the full distribution of Dice values; the denser, taller violin for USE shows both a higher median and a tighter clustering of scores. Collectively the six views demonstrate that coupling SAM2 with a rule-based expert system yields the most reliable boundaries and best volumetric agreement, whereas the LSTM variant exhibits larger errors and greater inter-patient variability.

**Figure 10.**
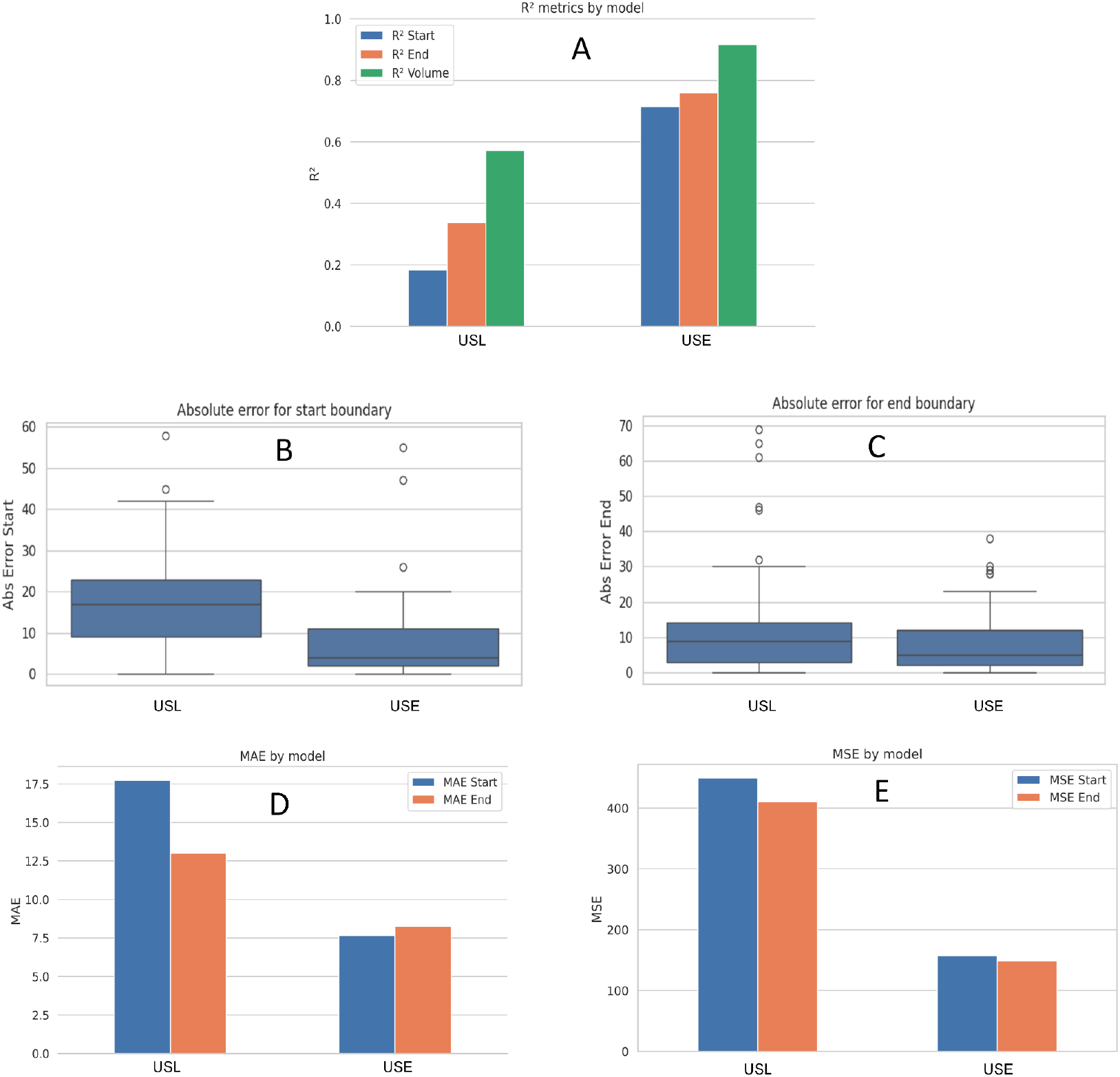
Performance comparison between the USL and USE pipelines. (A) Bar chart of R^2^ for start boundary, end boundary, and surrogate volume, showing how much variance in the reference labels each model explains. (B) and (C) Box and whisker plots of absolute error for start and end slice indices; shorter bars indicate finer localization. (D) Bar charts of Mean Absolute Error (MAE) for start and end boundary for each approach. (E) Bar charts of Mean Squared Error (MSE) for start and end boundary for each approach.

**Figure 11.**
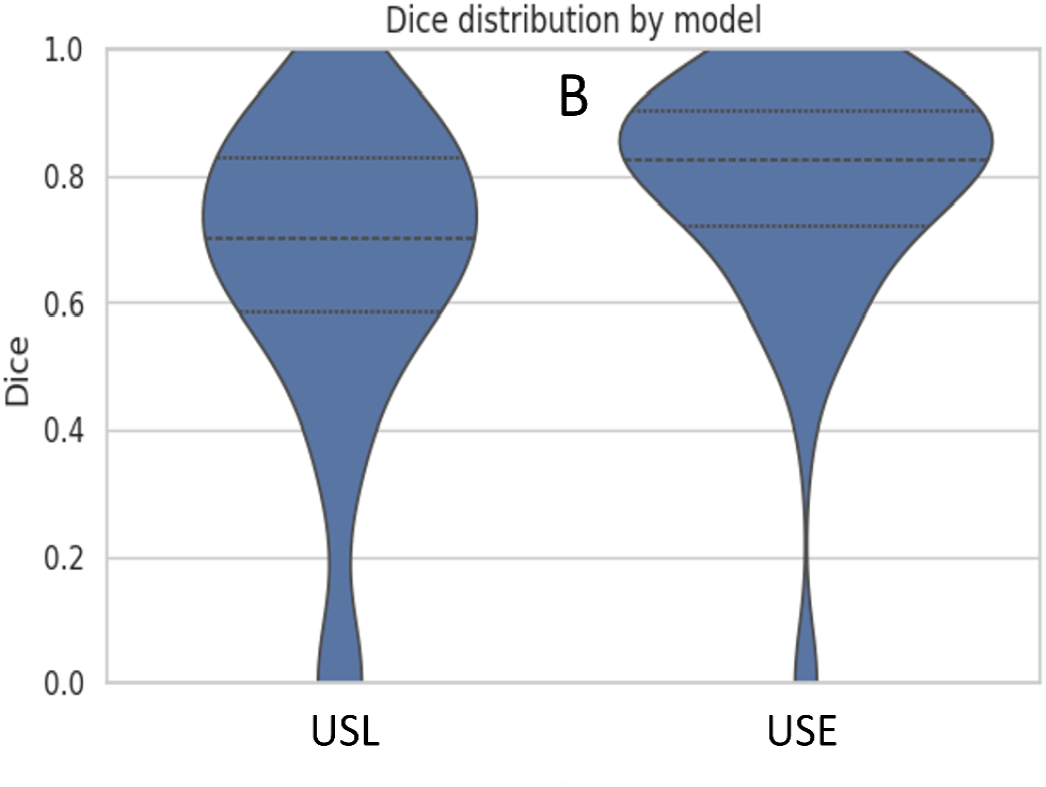
Violin plot of Dice distributions, revealing the full density of overlap scores for each pipeline.

## 5. Discussion

The side by side metrics confirm the clear advantage of the UNet + SAM2 + Expert (USE) pipeline over the UNet + SAM2 + LSTM (USL) alternative. For boundary localisation USE attains R^2^=0.714 for the start and 0.759 for the end, compared with only 0.183 and 0.340 for USL. That statistical edge translates into sharply lower localisation errors: the mean-absolute error falls from 17.8 to 7.7 at the start boundary and from 13.0 to 8.3 at the end, while the mean-squared error drops by roughly two-thirds (449→157 for the start, 410→149 for the end). Spatial overlap follows the same pattern, with the Dice coefficient rising from 0.64 for USL to 0.78 for USE. Most importantly for clinical use, surrogate lumen volume correlation climbs from R^2^ = 0.572 to 0.916, and the average volume error shrinks from 17% to about 11%. These results confirm that a deterministic sliding-window rule, when fed a high-quality SAM2 mask, can deliver reliable, interpretable boundary placement without the need for additional training data, a critical advantage in data-sparse clinical environments.

The strength of USE is especially relevant given the scarcity of labeled data. Our entire cohort comprises only 60 annotated samples. Such modest sample sizes restrict deep learning models with a high number of parameters, making a deterministic rule set an attractive choice. The LSTM was trained with conservative augmentation and a relatively deep recurrent stack, its performance therefore represents an early baseline rather than the final word on sequence learning. We anticipate that lighter recurrent blocks, positional embeddings, attention pooling, Dice-augmented loss, and channel stacking (raw signal, derivatives, smoothed counts) could greatly improve USL performance.

## 5. Conclusions

This study introduces two complementary pipelines that transform a UNet trained only on normal anatomy into a fully automated tool for abdominal aortic aneurysm (AAA) detection and quantification.

- Segmentation failure screening. The absence of a UNet mask in anomalous slices was turned into a binary classifier with a simple rolling-window rule. Tested on 33 scans (16 normal, 17 AAA), the method achieves 87.9 % accuracy, 88.2 % precision and recall, and an F1-score of 0.88, demonstrating that a label-free network can still act as a reliable triage filter.
- Multi-system quantification. Seeding Meta’s SAM2 with the UNet output enabled dense aortic masks through diseased segments. Two boundary-detection strategies were compared:
  ○ The UNet + SAM2 + Expert (USE) pipeline achieved R^2^ of 0.714 for start and 0.759 for end boundaries, MAE of 7.7 for start and 8.3 for end boundaries. Its Dice score was 0.78, and it yielded a strong surrogate volume R^2^ of 0.916.
  ○ The UNet + SAM2 + LSTM (USL) pipeline produced R^2^ of 0.183 for start and 0.340 for end boundaries, MAE of 17.8 for start and 13.0 for end boundaries. Its Dice score was 0.64, and the surrogate volume R^2^ of 0.572.

The deterministic expert rules therefore delivered markedly superior boundary localisation and volume agreement while requiring no additional training with R^2^ of 0.916. This work yields three key insights. First, when a UNet trained exclusively on normal aorta fails to produce a mask, that absence itself could serve as a marker of pathology. Second, even the sparse masks the UNet does generate are enough to “prompt” SAM2 into producing a continuous, high-quality aortic segmentation that spans the entire scan, aneurysm included. Third, a simple sliding-window rule applied to the SAM2 pixel-count signal can result in reliable aneurysm boundaries detection, achieving a volume correlation of roughly 0.92. Together these findings make the USE pipeline an end-to-end, interpretable solution for AAA screening and quantification in data-sparse settings, dramatically reducing manual annotation effort and opening a path toward scalable, fully automated vascular analysis. Important limitations remain: Volumes are derived from pixel-count surrogates rather than calibrated voxel data, and the current masks cover only the aneurysm lumen, not the outer wall or thrombus. Future work will integrate slice-thickness metadata, extend segmentation to the full vessel wall (including mural thrombus), and validate the approach on larger, multi-centre cohorts.

## Author Contributions

Conceptualization, S.M.L. and H.V.; methodology, A.B.R.; software, A.B.R.; validation, A.B.R. and B.P.; formal analysis, A.B.R., B.P., S.M.L. and H.V.; investigation, A.B.R., B.P., S.M.L. and H.V.; resources, S.M.L., H.V.; data curation, S.M.L. and B.P.; writing—original draft preparation, A.B.R.; writing—review and editing, S.M.L. and H.V.; visualization, A.B.R.; supervision, S.M.L. and H.V.; project administration, S.M.L. and H.V.; funding acquisition, S.M.L. and H.V. All authors have read and agreed to the published version of the manuscript.

## Institutional Review Board Statement

The data utilized in this study were sourced from the M2S Vascular Imaging Database, which provided fully de-identified data. Patient consent or Institutional Review Board (IRB) approval was not required, as the data were previously collected and anonymized by external organizations.

## Informed Consent Statement

Not required.

## Data Availability Statement

The data is available for purchase from M2S with an MOU. There is really no privacy concern, since it is de-identified data.

## Acknowledgments

We would like to thank Daniel Clair at Vanderbilt University as well as Shuvangee Dhar and Brian Haimerl for their help with this project.

## Disclaimer/Publisher’s Note

The statements, opinions and data contained in all publications are solely those of the individual author(s) and contributor(s) and not of MDPI and/or the editor(s). MDPI and/or the editor(s) disclaim responsibility for any injury to people or property resulting from any ideas, methods, instructions or products referred to in the content.

